# Protocol for the FIT FIRST Teen effectiveness and implementation study – a cluster randomised mixed methods trial

**DOI:** 10.1101/2025.08.01.25332617

**Authors:** Malte Nejst Larsen, Sofie Koch, Camilla Prisak Pedersen, Christina Birch Meiner, Chiara Cimenti, Caroline Eckert, Magnus Gjesing Christensen, Lars Breum Skov Christiansen, Cecilie Thøgersen-Ntoumani, Nikos Ntoumanis, Peter Krustrup, Giampiero Tarantino

## Abstract

**Introduction:** Physical activity (PA) is an important part of a healthy lifestyle for children and youth, but many adolescents do not fulfil the recommendations. The FIT FIRST concept can be a part of recommended PA, since it offers multiple sporting activities modified for the school setting, emphasizing high intensity, student engagement, enjoyment of movement, and inclusivity. The aim of the paper is to present the protocol for a study that will investigate the effects of FIT FIRST Teen over a full school year on cardiorespiratory fitness, body composition, muscle strength and quality of life as well as the fidelity, intervention dose, acceptability, appropriateness and feasibility of the intervention via an effectiveness-hybrid design (type 1).

**Methods and analysis:** A cluster-randomized controlled trial is conducted between August 2024 and August 2025 with 12 schools, >20 classes in an intervention group and 12 schools, >20 classes as controls. The effectiveness of the FIT FIRST Teen intervention will be evaluated through pre- and post-intervention testing over the course of one school year, using multi-level regression analysis to evaluate the intervention effects. Performance in Yo-Yo intermittent recovery level 1 test for children (YYIR1C) is the primary outcome. Secondary outcomes are blood pressure and resting heart rate, muscle strength, muscle mass, fat percentage, quality of life (including physical and psychological well-being, peer and social support, autonomy and parent relations, and school environment), body image, functionality appreciation, motivation for physical activity, sports club participation and self-perception.

To examine the potential for an impactful nationwide upscaling of the FIT FIRST Teen program, a comprehensive evaluation of the implementation will be conducted on all intervention schools. A mixed method approach will be adopted, with data collected via logbooks, observations, and interviews.

**Ethics and dissemination.:** The Regional Committees on Health Research Ethics for Southern Denmark (Videnskabsetisk Komité, Region Syd) has evaluated the FIT FIRST TEEN protocol and given it an ethics waiver (S-20210099). The study is registered at clinicaltrials.gov (NCT06615791) and the results will be disseminated through scientific papers and public engagement activities. Danish laws for collection and use of personal data will be followed, and parents will be asked for written content before publication.

## Introduction

Physical activity (PA) is an important part of a healthy lifestyle for children and youth, as it improves physical fitness, motor competencies and well-being, and thereby reduces the risk of developing overweight and non-communicable diseases later in life(1, 2). The Danish Health Authorities recommend that young people should participate in at least 60 minutes of moderate-to-vigorous PA (MVPA) per day, of which at least three times per week should involve muscle strengthening training(3). However, currently only about one-quarter of Danish 11-15-year-olds meet these recommendations(4). Self-determined motivation for physical activity is of interest as a long-lasting solution to this problem since it is a good predictor of sustained behaviour change(5).

Given that almost all children and adolescents spend a substantial portion of their waking hours in school, schools serve as pivotal settings for the widespread promotion of PA(6). However, current school-based PA interventions have shown limited effects on PA and fitness(7, 8). Often the poor results of interventions can be ascribed to implementation challenges, encompassing deficiencies in program fidelity, suboptimal attendance rates, or lack of access to intended resources, time, or space(9). Details about the contextual environment and how to implement interventions are typically derived from implementation evaluations. Nonetheless, such evaluations are rarely undertaken(10), but will be a focus point in the present study.

The FIT FIRST programme is built on the principles of using multiple sporting activities modified for the school setting, with a focus on high intensity, student engagement, enjoyment of movement, and inclusivity. A small-scale study has already shown great potential for the programme to increase physical health status(11). The use of multiple sporting activities modified for the school setting may also impact recess activities and leisure-time sport. Therefore, it will be investigated whether the developed abilities and motivation for FIT FIRST sporting activities can result in recruitment or re-recruitment of members to the local sports clubs.

The implementation of the FIT FIRST concept involves training teachers to deliver the programme, which has been developed by experienced staff from the University of Southern Denmark (SDU), the Danish Sport Confederation (Danmarks Idrætsforbund, DIF) and Team Denmark. These partners have developed three to six lessons for each of the included sports, all using pair-based exercises and small-sided game drills and activities. All the lessons included in the FIT FIRST Teen manual have been developed to provide high intensity cardiovascular and strength training, and to be motivating for children to participate, according to self-determination theory by securing the opportunity to fulfill the three basic psychological needs: autonomy, competences and relatedness(12).

### Objectives

This study aims to conduct a school-year-long cluster Randomised Controlled Trial (RCT) for adolescents in Denmark and assess 1) whether the FIT FIRST Teen intervention can improve adolescents’ physical fitness, motivation, sports club participation and well-being and 2) how the intervention is adopted and delivered across schools.

For the effect evaluation it is hypothesized that students from the schools randomised to the intervention arm will show greater improvements in cardiometabolic and muscular fitness, body composition, quality of life, body image outcomes, and motivation to engage in physical education (PE) than students randomised to the control arm.

For the implementation outcomes it is evaluated if the intervention is delivered with fidelity across all schools, and found acceptable, appropriate and feasible by the teachers involved. The mixed methods approach involving observations and interviews will provide understanding of the potential for improved implementation and scale-up. The implementation research will be conducted and reported in accordance with the requirements of the Standards for Reporting Implementation Studies (StaRI) Statement(13).

## Methods and analysis

### Trial Design

The study is a cluster-randomized controlled trial, with two arms and an allocation ratio of 1:1, using an effectiveness-hybrid design (type 1) testing for superiority of the intervention as well as exploratory factors. A SPIRIT schedule of enrollment, interventions, and assessments can be found in figure 1.

**Figure1:**
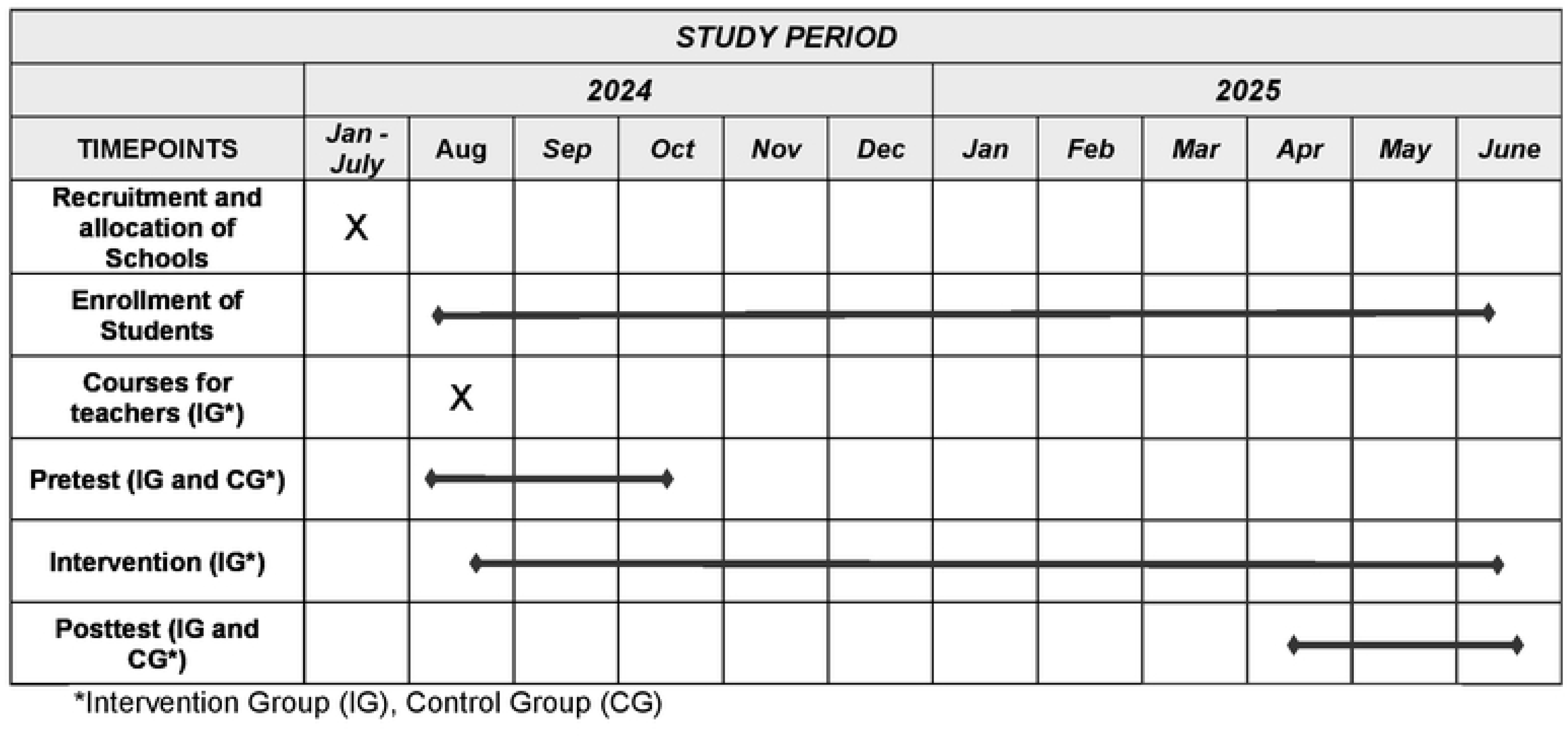
Timeline FIT FIRST Teen Study.

### Participants, interventions, and outcomes

#### Study setting

Compulsory schools in Denmark have been identified through websites of the Department of Education and local authorities. Posts on social medias (LinkedIn and Facebook) supported the recruitment efforts, and about 85% of the schools across all Danish regions have been invited by e-mails and calls. The proces can be followed in figure 2.

**Figure 2:**
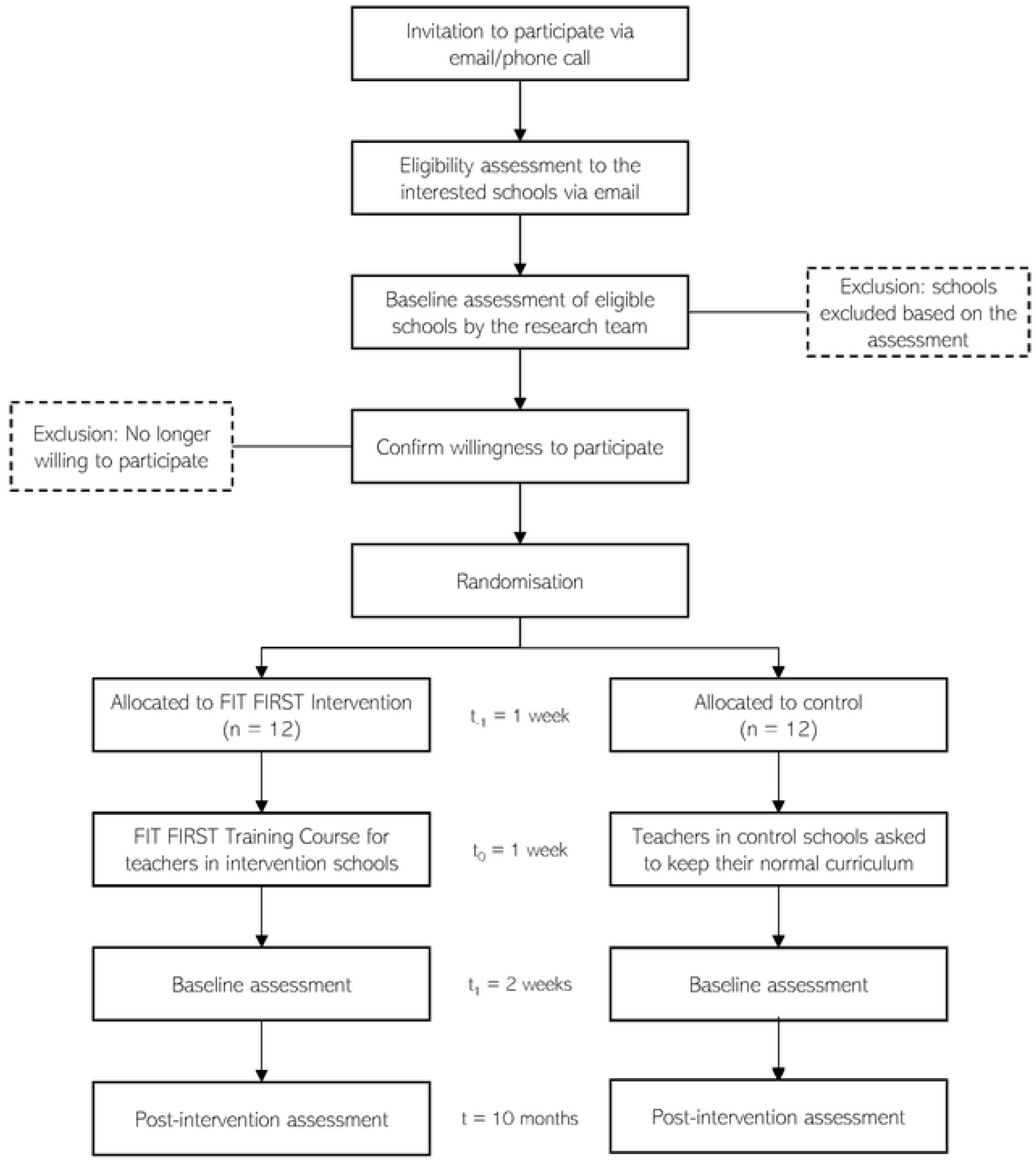
Recruitment chart.

#### Eligibility criteria

The schools were required to participate with at least two classes from 7^th^ to 9^th^ grade (11-to 16-year-olds) and were excluded if they already offer a special physical education program, e.g. elite sport classes. All pupils and teachers in the participating classes can be included in the study.

#### Intervention

Schools that agree to participate in the study will be randomised into two arms, intervention and control. The schools in the control arm will continue their normal PE-curriculum, and they will receive the intervention at the end of the trial.

Intervention arm: The intervention arm will implement three weekly 40-min FIT FIRST TEEN lessons over the course of nine months, corresponding to a full school year except the period before pre-testing and after post-testing (Fig. 1). The teachers who will teach these lessons will be invited to attend a one-day training course on the FIT FIRST Teen concept. This training course took place at the beginning of the school year 2024/25 (August-October 2024). The courses are comprised of theoretical sessions (training, physiology, and introduction to self-determination theory to foster motivation) of the FIT FIRST Teen concept as well as practice sessions where the teachers can experience the activities and discuss the content. The courses also focus on how to use the manual which outlines all the lessons to be implemented in the curriculum thrice a week over the full school year. Also, the schools will receive an equipment package with balls, bib jerseys and cones relevant to the activities.

The FIT FIRST Teen manual contains a total of 135 full 40-minute lesson descriptions, spread across 26 widely recognized leisure-time sports (Table 1). Each sport has been modified to consist mostly of pair-based exercises, small-sided games and relays with active rest, removing the most complex rules and sport specific techniques. Such adaptations maximize active time, allowing for students without prior experience with the sport in question to participate actively and feel competent, despite their lack of experience. Each lesson is designed to elicit an average heart rate (HR) of 75% of individual max HR, with at least 10% of the time being above 90% max HR. Such HRs have been shown in an unpublished study of ours, using similarly aged students who wore Polar Pro heart rate monitors. The sessions are designed with low equipment demands, and most of them can be performed both indoors and outside, minimizing logistic issues. The teachers have full autonomy in choosing what sports to perform over the course of the school year, as well as the scheduling, however they are encouraged to place the sessions on Mondays, Wednesdays and Fridays. One FIT FIRST session can replace the regular PE class, while the other two will be additional, but within regular school hours. The outlines of three full lessons translated into English can be found in supplementary material A).

**Table 1.** The 26 sports in the FIT FIRST Teen manual.

| **Sports in the FIT**<br>**FIRST Teen**<br>**manual** |  |
| --- | --- |
| **Sport** | **No. of lessons** |
| **Athletics** | **6** |
| **Automobile sports** | **6** |
| **Badminton** | **6** |
| **Basketball** | **6** |
| **Table tennis** | **6** |
| **Flag Football** | **6** |
| **Floorball** | **3** |
| **Soccer** | **6** |
| **Football tennis** | **3** |
| **Functional fitness** | **6** |
| **Handball** | **6** |
| **Field hockey** | **6** |
| **Judo** | **6** |
| **Ju-Jitsu** | **6** |
| **Canoe and kayak** | **6** |
| **Karate** | **6** |
| **Kickboxing** | **3** |
| **Rugby** | **6** |
| **Powerlifting** | **6** |
| **Squash** | **2** |
| **Taekwondo** | **6** |
| **Tennis** | **2** |
| **Padel tennis** | **2** |
| **Triathlon** | **6** |
| **Ultimate frisbee** | **6** |
| **Volleyball** | **6** |
| **Total:** | **135** |

Control arm: Teachers from the school in the control group will not receive the educational training nor the manual during the intervention phase. Before the baseline assessment, teachers in the control arm will be asked to continue their normal physical education curriculum for the entire duration of the trial. However, after the end of the study, teachers in the control arm will be offered the same educational training and manual as an incentive to participate in the present study.

### Participant timeline

Enrolment and pre-intervention data collection will take place at the beginning of the school year 2024/25 (August-October 2024). Follow-up data collection will take place during May and June 2025, and the process evaluation will be finalized in August 2025. An overview of the data collection can be found in table 2.

**Table 2.** Summary of measures used in the FIT FIRST Teen study and time points.

|  | Measurement instruments | Baseline | Follow-up |
| --- | --- | --- | --- |
| **Objective measures** |  | X | X |
| Height | Tanita stadiometer | X | X |
| Body composition | Inbody 270 bioimpedance(15) | X | X |
| Cardiometabolic fitness | Yo-Yo Intermittent Recovery level 1<br>Children’s test(14) | X | X |
| Muscular fitness | Counter-movement jump, arrowhead agility,<br>stork balance and handgrip strength (16-18) | X | X |
| Resting heart rate and systolic<br>and diastolic blood pressure | Oscillometry | X | X |
| **Self-reported measures** |  |  |  |
| Fidelity | The Conceptual framework for implementation fidelity(36). | X | X |
| Intervention dose | The Conceptual framework for implementation fidelity(36). | X | X |
| Acceptability, Appropriateness, Feasibility | The Acceptability of Implementation Measure, Implementation Appropriate Measure, and Feasibility of Intervention Measure<sup>31</sup> | X | X |
| Motivation for physical activity | Perceived Locus of Causality Scale | X | X |
| Quality of life | KIDSCREEN-27 | X | X |
| Well-being | Physical and psychological well-being domains in the KIDSCREEN-27 instrument | X | X |
| Body image | Body Appreciation Scale-2 (BAS-2) | X | X |
| Physical self-perception | Physical Self-Inventory (PSI-SF) | X | X |
| Demographics | Age and ethnicity | X |  |

### Outcomes

All quantitative outcomes will be aggregated by mean and standard deviation scores.

#### Primary outcome

Cardiorespiratory fitness: The Yo-Yo Intermittent Recovery Level 1 test for children (YYIR1C) will be conducted to evaluate cardiorespiratory fitness levels(14). The participants will run 16 meters back and forward with a 10 seconds pause after every 2×16 m shuttle run. They will follow an increasing pace by beep sounds, until they cannot reach the finish line in time. In such a case, the first time they will be warned, but in the second time the test will end, and the reached level will be noted by testing personnel.

Also resting heart rate and blood pressure measurement will be measured at rest in a supine position with Welch Allyn ProBP 3400 devices. Three measurements will be conducted after an 8-minute resting period.

#### Secondary outcomes

Body composition: Tanita Height scales and InBody 270 bioimpedance measurements will be taken to assess body mass, lean mass, fat mass, fat percentage and body mass index (BMI) z-score, prevalence of overweight and obesity(15).

Muscular fitness: To evaluate the intervention effects on muscular fitness, the horizontal countermovement jump test (best result of two attempts), the arrowhead agility test (best result of two attempts), the stork balance test (best result of three attempts standing on each of the legs) and the handgrip strength test will be used (best result of two attempts)(16–18).

To investigate the potential impact of the FIT FIRST Teen program on students’ well-being, motivation, sports club participation and body perceptions, a series of questionnaires will be used.

Health-Related Quality of Life: The Health-Related Quality of Life (HRQoL) will be assessed using the KIDSCREEN-27 (KS-27)(19). This instrument has been widely used and validated in the Danish context(20, 21) (22). The KIDSCREEN-27 domains include physical well-being (5 items), psychological well-being (7 items), peer and social support (4 items), autonomy and parent relations (7 items), and school environment (4 items). A 5-point Likert scale, ranging from 1 to 5, will be used. Depending on the item, the value labels will vary between 1, “*not at all*” to 5, “*extremely*” and 1, “*never*” to 5, “*always*”.

Body image: Body image outcomes will be operationalized in terms of body appreciation and functionality appreciation. Body appreciation has been defined as the ability to accept, hold favorable opinions toward, and respect the body, while also rejecting media-promoted appearance ideals as the only form of human beauty(23). To investigate adolescents’ body appreciation, the 10-item Body Appreciation Scale-2 (BAS-2) will be used(24). This one-dimension instrument has been validated among Danish adolescents(25). A 5-point Likert scale, ranging from 1, “*Never”,* to 5, “*Always”*, will be used. Functionality appreciation is defined as the “appreciation of, respect for, and honoring one’s body for what it can do and is capable of doing”(24). The 7-item Functionality Appreciation Scale (FAS) will be used to measure this outcome(26). Such an instrument comprises seven items and has been validated in different cohorts of adolescents(27, 28). The items will be accompanied by a 5-point Likert scale, ranging from 1, “*strongly disagree”,* to 5, “*strongly agree”*.

Motivation for physical activity: The Perceived Locus of Causality Scale(*29*) will be used to examine students’ motivational regulations towards PE. Based on SDT(30), the scale has four items to measure intrinsic motivation, identified regulation, introjected and external regulation and Ntoumanis has added an amotivation subscale. Therefore, the 20-item scale developed by Ntoumanis(31) will be employed to assess students’ motivation towards PE. The scale will be accompanied by a 7-point Likert scale ranging from 1, *“strongly disagree”,* to 7, “s*trongly agree”*. Moreover, the participants will be asked if they participate in leisure time sports activities.

Physical self-perception: To investigate physical self-perception, defined as a multidimensional and hierarchical model that comprises subdomains like sport competence, physical condition, body attractiveness, and physical strength(32), the short form of the Physical Self-Inventory (PSI-SF) will be used(33). This 18-item inventory comprises items that measure global self-concept, physical self-worth, physical condition, sports competence, physical attractiveness, and physical strength(34). A 6-point Likert scale, ranging from 1, “*not at all”,* to 6, *“entirely”,* will accompany each item. The PSI-SF has been widely used and validated in adolescent populations across different countries(34, 35).(32–35)

Fidelity and dose delivered/received: To examine the degree to which the FIT FIRST Teen program is implemented as intended, the Conceptual framework for implementation fidelity will be used(36). The framework includes a list of factors that cover fidelity (e.g., content, duration, complexity), but also can be used as a measure for intervention dose (e.g., duration, frequency, participant responsiveness).

Acceptability, appropriateness and feasibility: The Acceptability of Intervention Measure, Intervention Appropriateness Measure, and Feasibility of Intervention Measure(37) will be used to measure acceptability, appropriateness and feasibility of the FIT FIRST Teen program. The questionnaire contains four questions for each of the three constructs (acceptability, appropriateness, feasibility), and employs a 5-point Likert scale, ranging from 1, “Completely disagree”, to 5, “Completely agree”.

### Sample Size

A priori power analysis was conducted using the Cluster-Randomized Trials calculator by the National Institute of Health [Accessed on 20/03/2024] (Available from: https://researchmethodsresources.nih.gov/.) to determine the minimum cluster size required to test the study hypotheses. The calculation is based on a previous meta-analysis that investigated the differences in adolescents’ cardiorespiratory fitness after a school-based intervention, which reported a median effect size of *d =* 0.26(38). Statistical power was set at .90 and α = .05. Moreover, we assumed on average 19 participants per cluster (class) with expected correlations over time at a member-level of 0.87 and at a cluster-level of 0.498, using pilot data from an application of FIT FIRST in the Faroe Islands(11). Finally, using the same data, proportion of variance at the member- and cluster-levels were set at 0.1011 and 0.062, respectively(11). Based on these parameter estimates, the results from the power analysis indicated that the number of clusters (classes) needed per arm was 20. However, 20% more classes were added per condition to account for possible attrition. Therefore, the final sample size consisted of 48 classes, which resulted in 10 – 12 schools (3-4 classes per school), and 912 students in total.

### Recruitment Strategies

To ensure adequate enrolment to achieve the target sample size, different recruitment strategies will be employed. All schools in Denmark have been invited to participate in the study by social medias and schools in Seeland and Funen has also been invited by e-mails and phone calls. All pupils and parents will be informed of the study through the schools’ communication platforms and can decide whether they want to participate or not.

### Assignment of Interventions

Allocation: Once schools are recruited, they will be assigned to either control or intervention arm, with a 1:1 allocation, employing a code generated using the Survey Sampling R package version 2.10 in RStudio(39). The randomization will be stratified by number of classes per school, and school location (namely, rural vs urban areas) to ensure equal sample size and location representation in each group.

Blinding: Considering the nature of the intervention, it is not possible to blind experimental allocation to researchers delivering the FIT FIRST training to teachers, as well as outcome assessors. However, blinding procedures will be employed for the data analyst. Specifically, to ensure blinding, the data analyst will receive datasets in which the experimental groups are coded anonymously (e.g., labeled simply as Group A, Group B). The allocation key linking anonymized group labels to actual intervention conditions will be withheld until after all primary statistical analyses are completed, thereby preventing potential bias during data interpretation.

### Data Collection, Management and Analysis

Data collection methods: Data will be collected using validated instruments, questionnaires and fitness tests. The data will be collected at baseline (at the beginning of the school year, fall 2024) and at a 10-month follow-up (at the end of the school year, spring 2025) by research assistants who will be trained to collect such data. They will visit each class one planned day for each test round, testing each class at the schools’ own facilities. We will encourage all participants to be a part of the follow-up assessment, but it is voluntary, and if they are not available on the planned test day, no measures will be taken.

To examine the implementation of the FIT FIRST Teen program, a comprehensive evaluation of the implementation will be conducted. For the purpose of complementarity, a mixed-methods approach will be adopted, including logbooks, observations, and interviews(40).

Logbooks: Each class will be provided with a logbook, which the teacher(s) involved will be responsible for completing on a weekly basis. The logbook consists of questions regarding the weekly number of FIT FIRST Teen sessions, days of FIT FIRST Teen delivery, and the sport practiced each week. The aim of the logbook is to measure FIT FIRST Teen fidelity and intervention dose.

Questionnaires: Teachers participating in the delivery of FIT FIRST at all intervention schools will receive a short questionnaire twice during the intervention period. The aim of the questionnaire is to collect data on intervention acceptability, appropriateness, and feasibility, and will be guided by the Acceptability of Intervention Measure (AIM), Intervention Appropriateness Measure (IAM), and Feasibility of Intervention Measure (FIM)(37). (41), and relevant background information (e.g., physical education background).

Observations: Observations will be conducted three times (start (fall 2024), mid-way (winter 24/25, end (summer 2025) at six selected intervention schools. At each school one class, including one or two teachers, will be subject for observation. At each of the three observations point, a full FIT FIRST session will be observed. The observations will be conducted at the same weekday at each observation timepoint. The observations will be guided by the Consolidated Framework for Implementation Fidelity (CFIF) (36)a(42) Specifically, CFIF will assess intervention adherence (e.g., to what extent do they follow the manual), intervention dosage/exposure (e.g., time allocated to FIT FIRST), coverage (e.g., percentage of students that participate), quality (e.g., to what extent the FIT FIRST session focus on high intensity, inclusion of all students and fun), participant responsiveness (e.g., student engagement), program differentiation (e.g., key elements linked to positive outcomes), intervention complexity (e.g., students understanding and clarity of FIT FIRST activities), and facilitation strategies (e.g., teacher strategies to enhance fidelity). The aim of the observations is to gain insight into practice and the execution of the FIT FIRST TEEN sessions. Selection of schools will be based on the following criteria: geographic location (western/eastern part of Denmark), school size (large/small), urban/rural location, and public/private schools.

Interviews: Single interviews will be conducted twice during the intervention period at the six schools that are also selected for observation, with one teacher per school, who is responsible for the FIT FIRST TEEN delivery. The interviews will be guided by Consolidated Framework for Implementation Research (CFIR), which is one of the most widely used frameworks to guide assessment of contextual determinants of implementation (42) (42, 43) Specifically, the interviews will focus on the following domains CFIR domains: innovation domain (e.g., innovation adaptability and complexity), inner setting (e.g., structural characteristics, culture, relative priority, available resources), individuals domain (e.g., implementation leads, innovation delivers, innovation recipients), characteristics subdomain (e.g., capability, opportunity, and motivation), and implementation process (e.g., assessing needs, planning, engaging, adapting). The interviews will also focus on the three implementation outcomes highlighted by Proctor et al. (44); acceptability, appropriateness, and feasibility. The aim of the interviews is to gain a deeper understanding of the translation of FIT FIRST into school practice. Additionally, the aim is to examine how FIT FIRST is organized at the schools and to elucidate the facilitators and barriers to the implementation of FIT FIRST program in the lower secondary school.

#### Data Management

All data from the objective measures will be written on papers on the testing days, and thereafter noted in an excel database. The papers will be stored according to the rules for data storage. All data from the questionnaires will be stored in SurveyXact. All observation notes will be written on papers during each observation. Interviews will be recorded using a voice recorder, and thereafter transcribed verbatim into a word document. All papers, observation notes, voice record files, and interview transcriptions will be stored according to the rules for data storage. All data from the questionnaires will be stored in SurveyXact.

#### Statistical Methods

Descriptive analysis will be carried out to report means and standard deviations of primary and co-primary outcomes for each arm at baseline and 10-month follow-up. To examine the effectiveness of the intervention, mixed linear (multilevel) modelling will be employed to examine changes across and between arms in all measures over time, adjusting for clustering effects and baseline measures of the outcome variables. Intention-to-treat analysis will be used to deal with missing data and will compare these results against an analysis using all available data (with the full information maximum likelihood method). Sensitivity analysis will be conducted to examine the prognostic effects of age, height and sports club participation to compare the parameter estimates of the models with and without the covariates. Trial fidelity, dose, acceptability, appropriateness, and feasibility will be analysed using one sample t-tests, examining potential differences from pre to post intervention. Descriptive statistics and tables will be used to visualize data collected from logbooks and teacher questionnaires.

Qualitative data will be audio-recorded and transcribed verbatim and uploaded into NVivo software for analysis (45). A framework thematic analysis approach will be used to code and interpret the data for the aim of exploring the acceptability, appropriateness, feasibility, and implementation fidelity of the FIT FIRST Teen intervention, including key facilitators and barriers. The framework thematic analysis consists of five iterative phases: Familiarization, development of the codebook, indixing, charting, and interpretation (46–48). The analysis will be conducted using the CFIF and CFIR frameworks. Observational data will be used to support and triangulate findings (e.g., by providing contextual examples or elaborating on interview-derived themes).

### Strengths and limitations of this study

- The nationwide sample will lower the risk for selection bias.
- The examination of a broad selection of health outcomes will test intervention efficacy for physical as well as mental health.
- The mixed method approach will give a detailed insight into the potential impact of the intervention.
- Due to practical constraints with field testing at schools, it is not possible to administer gold standard measures of body composition and fitness.
- Lack of data about socioeconomic factors as potential confounding factors is a limitation.

### (49)The Patient and Public Involvement statement

None

### Ethics and Dissemination

The Regional Committees on Health Research Ethics for Southern Denmark (Videnskabsetisk Komité, Region Syd) has evaluated the FIT FIRST Teen protocol and given it an ethics waiver (S-20210099, supplementary material B). The study is registered at clinicaltrials.gov (NCT06615791).

Any protocol modifications will be reported when results are published and any deviations from the protocol will also be reported in the trial registry.

Informed consent will be obtained from the children’s parents or guardians before publication of the results, according to journal guidelines. In addition, informed consent will be obtained from teachers participating in interviews. At baseline, each participant will be assigned a random alphanumeric identifier and data will be anonymized after collection to maintain confidentiality. All participants will be informed that participation is voluntary, and that they can withdraw at any time.

Throughout the duration of the intervention, all study-related data will be stored in encrypted OneDrive and SharePoint documents. All Principal Investigators and the research team of the FIT FIRST Teen project will be given access to the datasets. Sharing of the data will happen according to Danish rules.

The pre-registered primary and co-primary outcomes of the trial will be reported following the CONSORT statement for cluster RCTs and published in peer-reviewed journals. Results will also be presented at academic conferences. Finally, lay summaries will be provided to the participating schools, local stakeholders, and parents of the participants on the project’s website that will be created. Authors and contributors to publications related to the project will be defined following the recommendations of the International Committee of Medical Journal Editors (ICMJE).

## Data Availability

No datasets were generated or analysed during the current study. All relevant data from this study will be made available upon study completion.

## Data statement

Data will be available upon request (send e-mail to corresponding author) within the Danish laws of data sharing.

## Author Contributions

MNL, SK, MGC, GT, CBM, CE, CC, CPP, NN, LBSC, CT and PK have been involved in the design of the study. MNL, NN, LBSC, CT and PK contributed to the funding application with PK as the main applicant. MNL, SK and GT have written the first draft of the manuscript. MNL, SK, MGC, GT, CBM, CE, CC, CPP, NN, LBSC, CT and PK have been part of the revision and have approved the final draft of the manuscript. MNL is the guarantor.

---

Figure 1: Flow Chart

## References

1. Bangsbo J, Krustrup P, Duda J, Hillman C, Andersen LB, Weiss M, et al. The Copenhagen Consensus Conference 2016: children, youth, and physical activity in schools and during leisure time. British journal of sports medicine. 2016;50(19):1177–8.

2. Putra IGNE, Daly M, Sutin A, Steptoe A, Scholes S, Robinson E. Obesity, psychological well-being related measures, and risk of seven non-communicable diseases: evidence from longitudinal studies of UK and US older adults. International Journal of Obesity. 2024:1–9.

3. Toftager M, Brønd JC, Roman JEI, Kristensen PL, Damsgaard MT, Grøntved A, et al. Måling af fysisk aktivitet i Skolebørnsundersøgelsen. 2024.

4. Steene-Johannessen J, Anderssen SA, Kolle E, Hansen BH, Bratteteig M, Dalhaug EM, et al. Temporal trends in physical activity levels across more than a decade–a national physical activity surveillance system among Norwegian children and adolescents. International Journal of Behavioral Nutrition and Physical Activity. 2021;18(1):55.

5. Ntoumanis N, Ng JY, Prestwich A, Quested E, Hancox JE, Thøgersen-Ntoumani C, et al. A meta-analysis of self-determination theory-informed intervention studies in the health domain: Effects on motivation, health behavior, physical, and psychological health. Health psychology review. 2021;15(2):214–44.

6. Dobbins M, Husson H, DeCorby K, LaRocca RL. School-based physical activity programs for promoting physical activity and fitness in children and adolescents aged 6 to 18. Cochrane database of systematic reviews. 2013(2).

7. Neil-Sztramko SE, Caldwell H, Dobbins M. School-based physical activity programs for promoting physical activity and fitness in children and adolescents aged 6 to 18. Cochrane database of systematic reviews. 2021(9).

8. Jones M, Defever E, Letsinger A, Steele J, Mackintosh KA. A mixed-studies systematic review and meta-analysis of school-based interventions to promote physical activity and/or reduce sedentary time in children. Journal of Sport and Health Science. 2020;9(1):3–17.

9. Jago R, Salway R, Beets M, Lubans DR, de Vocht F. Rethinking children’s physical activity interventions at school: a new context-specific approach. Frontiers in Public Health. 2023;11:1149883.

10. Durlak JA, DuPre EP. Implementation matters: A review of research on the influence of implementation on program outcomes and the factors affecting implementation. American journal of community psychology. 2008;41:327–50.

11. Olsen HW, Sjúrðarson T, Danielsen BB, Krustrup P, Larsen MN, Skoradal M-B, et al. A 10-week implementation of the FIT FIRST FOR ALL school-based physical activity concept effectively improves cardiorespiratory fitness and body composition in 7–16-year-old schoolchildren. Frontiers in Public Health. 2024;12:1419824.

12. Ryan RM, Deci EL. Intrinsic and extrinsic motivation from a self-determination theory perspective: Definitions, theory, practices, and future directions. Contemporary educational psychology. 2020;61:101860.

13. Pinnock H, Barwick M, Carpenter CR, Eldridge S, Grandes G, Griffiths CJ, et al. Standards for reporting implementation studies (StaRI) statement. bmj. 2017;356.

14. Bendiksen M, Williams CA, Hornstrup T, Clausen H, Kloppenborg J, Shumikhin D, et al. Heart rate response and fitness effects of various types of physical education for 8-to 9-year-old schoolchildren. European journal of sport science. 2014;14(8):861–9.

15. Larsen MN, Krustrup P, Araújo Póvoas SC, Castagna C. Accuracy and reliability of the InBody 270 multi-frequency body composition analyser in 10-12-year-old children. PLoS One. 2021;16(3):e0247362.

16. Tomkinson GR, Carver KD, Atkinson F, Daniell ND, Lewis LK, Fitzgerald JS, et al. European normative values for physical fitness in children and adolescents aged 9–17 years: results from 2 779 165 Eurofit performances representing 30 countries. British journal of sports medicine. 2018;52(22):1445–56.

17. Kranti Panta B. A study to associate the Flamingo Test and the Stork Test in measuring static balance on healthy adults. Foot Ankle Online J. 2015;8(4).

18. Rago V, Brito J, Figueiredo P, Ermidis G, Barreira D, Rebelo A. The arrowhead agility test: reliability, minimum detectable change, and practical applications in soccer players. The Journal of Strength & Conditioning Research. 2020;34(2):483–94.

19. Ravens-Sieberer U, Auquier P, Erhart M, Gosch A, Rajmil L, Bruil J, et al. The KIDSCREEN-27 quality of life measure for children and adolescents: psychometric results from a cross-cultural survey in 13 European countries. Quality of Life Research. 2007;16(8):1347–56.

20. Madsen M, Elbe AM, Madsen EE, Ermidis G, Ryom K, Wikman JM, et al. The “11 for Health in Denmark” intervention in 10-to 12-year-old Danish girls and boys and its effects on well-being—A large-scale cluster RCT. Scandinavian Journal of Medicine & Science in Sports. 2020;30(9):1787–95.

21. Nielsen T. Validering af den danske udgave af KIDSCREEN-27 børn og unge selv-rapporteringsversion. Psykometriske egenskaber undersøgt ved hjælp af Rasch-analyser: Projektet” Validering af KIDSCREEN-27 (Tillægsundersøgelse ifm. Børns Vilkårs undersøgelse af skolebørns livskvalitet)” er gennemført af Tine Nielsen, UCL Erhvervsakademi og Professionshøjskole, Afdelingen for Anvendt Forskning i Pædagogik og Samfund, med datagrundlag fra Børns Vilkår og fondsstøtte fra Ole Kirk’s Fond. 2023.

22. Madsen M, Larsen MN, Cyril R, Møller TK, Madsen EE, Ørntoft C, et al. Well-Being, Physical Fitness, and Health Profile of 2,203 Danish Girls Aged 10–12 in Relation to Leisure-time Sports Club Activity—With Special Emphasis on the Five Most Popular Sports. Journal of Strength and Conditioning Research. 2022;36(8):2283–90.

23. Avalos L, Tylka TL, Wood-Barcalow N. The body appreciation scale: Development and psychometric evaluation. Body image. 2005;2(3):285–97.

24. Tylka TL, Wood-Barcalow NL. The Body Appreciation Scale-2: item refinement and psychometric evaluation. Body image. 2015;12:53–67.

25. Lemoine J, Konradsen H, Jensen AL, Roland-Levy C, Ny P, Khalaf A, et al. Factor structure and psychometric properties of the Body Appreciation Scale-2 among adolescents and young adults in Danish, Portuguese, and Swedish. Body image. 2018;26:1–9.

26. Alleva JM, Tylka TL, Van Diest AMK. The Functionality Appreciation Scale (FAS): Development and psychometric evaluation in US community women and men. Body image. 2017;23:28–44.

27. Sahlan RN, Todd J, Swami V. Psychometric properties of a Farsi translation of the Functionality Appreciation Scale (FAS) in Iranian adolescents. Body Image. 2022;41:163–71.

28. He J, Cui T, Barnhart WR, Chen G. The Chinese version of the Functionality Appreciation Scale: Psychometric properties and measurement invariance across gender and age. Journal of Eating Disorders. 2023;11(1):99.

29. Goudas M, Biddle S, Fox K. Perceived locus of causality, goal orientations, and perceived competence in school physical education classes. British Journal of Educational Psychology. 1994;64(3):453–63.

30. Ryan RM, Deci EL. Self-determination theory: Basic psychological needs in motivation, development, and wellness: Guilford publications; 2017.

31. Ntoumanis N. A self-determination approach to the understanding of motivation in physical education. British journal of educational psychology. 2001;71(2):225–42.

32. Fox KR, Corbin CB. The physical self-perception profile: Devlopment and preliminary validation. Journal of sport and Exercise Psychology. 1989;11(4):408–30.

33. Morin AJ, Maiano C. Cross-validation of the short form of the physical self-inventory (PSI-S) using exploratory structural equation modeling (ESEM). Psychology of Sport and Exercise. 2011;12(5):540–54.

34. Maïano C, Morin AJ, Ninot G, Monthuy-Blanc J, Stephan Y, Florent J-F, et al. A short and very short form of the physical self-inventory for adolescents: Development and factor validity. Psychology of Sport and Exercise. 2008;9(6):830–47.

35. Aşçi FH, Maïano C, Morin AJ, Çağlar E, Bilgili N. Validity and reliability of the Very Short form of the Physical Self-Inventory among Turkish adolescents. Journal of Sports Sciences. 2017;35(21):2060–6.

36. Carroll C, Patterson M, Wood S, Booth A, Rick J, Balain S. A conceptual framework for implementation fidelity. Implementation science. 2007;2:1–9.

37. Weiner BJ, Lewis CC, Stanick C, Powell BJ, Dorsey CN, Clary AS, et al. Psychometric assessment of three newly developed implementation outcome measures. Implementation science. 2017;12:1–12.

38. Minatto G, Barbosa Filho VC, Berria J, Petroski EL. School-based interventions to improve cardiorespiratory fitness in adolescents: systematic review with meta-analysis. Sports medicine. 2016;46:1273–92.

39. Tillé Y, Matei A. Sampling: survey sampling. R package version. 2009;2.

40. Greene JC. Mixed methods in social inquiry: John Wiley & Sons; 2007.

41. Huijg JM, Gebhardt WA, Crone MR, Dusseldorp E, Presseau J. Discriminant content validity of a theoretical domains framework questionnaire for use in implementation research. Implementation Science. 2014;9:1–16.

42. Damschroder LJ, Reardon CM, Opra Widerquist MA, Lowery J. Conceptualizing outcomes for use with the Consolidated Framework for Implementation Research (CFIR): the CFIR Outcomes Addendum. Implementation science. 2022;17(1):7.

43. Michie S, Van Stralen MM, West R. The behaviour change wheel: a new method for characterising and designing behaviour change interventions. Implementation science. 2011;6:1–12.

44. Proctor E, Silmere H, Raghavan R, Hovmand P, Aarons G, Bunger A, et al. Outcomes for implementation research: conceptual distinctions, measurement challenges, and research agenda. Administration and policy in mental health and mental health services research. 2011;38:65–76.

45. Jackson K, Bazeley P. Qualitative data analysis with NVivo. 2019.

46. Gale NK, Heath G, Cameron E, Rashid S, Redwood S. Using the framework method for the analysis of qualitative data in multi-disciplinary health research. BMC medical research methodology. 2013;13:1–8.

47. Parkinson S, Eatough V, Holmes J, Stapley E, Midgley N. Framework analysis: a worked example of a study exploring young people’s experiences of depression. Qualitative research in psychology. 2016;13(2):109–29.

48. Bryman A, Burgess RG. Analyzing qualitative data: Routledge London; 1994.

49. Braun V, Clarke V. Using thematic analysis in psychology. Qualitative research in psychology. 2006;3(2):77–101.

